# Effects of recreational sports and combined training on blood pressure and glycosylated hemoglobin in middle-aged and older adults: a systematic review and meta-analysis

**DOI:** 10.1101/2021.05.05.21256401

**Authors:** Vinícius Mallmann Schneider, Paula Frank, Sandra C. Fuchs, Rodrigo Ferrari

## Abstract

**Background:** Combined resistance and aerobic training (CT) is the most suitable form of exercise training to simultaneously improve cardiometabolic profile and functional capacity in middle-aged and older adults. Recreational sports (RS) emerge as an alternative to traditional exercises to improve these outcomes that could be used as a retention and continuity strategy, promoting health benefits associated with pleasure and satisfaction during the physical activity.

**Objectives:** The aim was to conduct a meta-analysis on the effects of RS and CT on systolic blood pressure (SBP), diastolic blood pressure (DBP) and glycosylated hemoglobin (HbA1c) in middle-aged and older adults and to compare these exercise interventions to a non-exercising control group (CON).

**Data Sources:** A literature search was conducted using the databases at PubMed, COCHRANE and SciELO between July and August 2020.

**Study Eligibility Criteria:** Studies that included men and women aged ≥45 years, healthy or with values of baseline for SBP ≥130mmHg or DBP ≥80 mmHg or with type II diabetes, in which the participants performed RS or CT versus CON, and evaluated SBP, DBP and HbA1c.

**Study Appraisal and Synthesis Methods:** Two independent reviewers screened search results, performed data extraction, and assessed of methodological quality of studies. Random effects modeling was used to compare pre-to post-intervention changes in BP and HbA1c from RS and CT versus CON, and the effect size were calculated through the weighted mean difference (MD) with a 95% confidence interval (CI).

**Results:** From 6017 records, 27 studies were included (9 RS and 18 CT interventions). The analysis included 1411 participants with 55 ± 8 years (range, 45–73 years). Overall, RS was associated reduction in SBP (MD: -7.20 mmHg; CI: -13.7 to -0.7) and DBP (MD: -3.6 mmHg; 95%CI: -6.6 to -0.5) versus CON. CT reduction SBP (MD: -3.6 mmHg; 95%CI: -5.2 to -2.0) and DBP (MD: -3.10 mmHg; 95%CI: -3.75 to -2.44) versus CON. RS not reduced HbA1c (MD: -0.24 %; CI: -0.53 to 0.06) versus CON. However, CT reduced HbA1c (MD: -0.47%; 95%CI: -0.67 to -0.27) versus CON.

**Conclusions:** RS and CT are effective exercise interventions to improve blood pressure in middle-aged and older adults. Additionally, CT seems to be an excellent strategy to reduce HbA1c, and future studies are necessary to confirm the effectiveness of recreational sports to improve HbA1c.

**Systematic Review Registration Number:** CRD42020207250

**Key Points:** Recreational sports promote similar or even greater blood pressure reduction when compared to traditional exercises.

Combined training promotes improvements in blood pressure and HbA1c in aging adults.

As alternative to traditional exercises, recreational sports emerge as an effective strategy to improve the cardiometabolic profile in middle-aged and older adults.

## 1. INTRODUCTION

Aging is a continuous phenomenon that leads to a progressive decline in physiological systems (cardiovascular, metabolic, and skeletal muscle), prone to development of noncommunicable diseases, such as hypertension, type II diabetes and coronary artery disease [1, 2]. Although this physiological decline is inexorable, there are alternatives to postpone its progression. Physical exercise is the cornerstone non-pharmacological intervention to improve functionally and cardiometabolic risk factors such as blood pressure (BP) and glycosylated hemoglobin (HbA1c) [3, 4]. Reductions of BP and glycemic level are key components to prevent and treat hypertension and diabetes, and different exercises can be effective for BP control and to lower HbA1c levels [3, 5].

The combination of strength and aerobic training (i.e., combined training) is among the most effective strategies to simultaneously improve physical fitness and prevent age-associated diseases [6, 7]. Although the benefits of combined training have been well described [8], individuals must consistently adhere to the practice to maintain the improvements. Environmental and physical barriers, in addition to lack of motivation resulted from the monotony of the traditional exercises can lead to low adherence to perform exercise regularly, and middle-aged and older adults are particularly vulnerable to failure [9]. Recreational sports emerges as an interesting alternative that could be used as a retention and continuity strategy, promoting health benefits associated with pleasure and satisfaction during the physical activity [10, 11]. The popularity of recreational sports in different countries, the easy accessibility to sports facilities and public areas to practice are among some potential advantages if compared to most traditional exercises. However, in middle-aged and older adults, scarcity data assessing the effects of recreational sports on blood pressure and glycemic profile is available [12], reinforcing the importance of new studies related to this topic.

In relation to the effectiveness of recreational sports and traditional exercises for BP and HbA1c, previous reviews have focused on general population, but no previous meta-analysis has assessed the effects of these two types of physical activity in older populations. Since combined training is the most recommended modality to counteract several age-related declines and recreational sports can also be effective to improve the cardiometabolic profile in aging adults, this systematic review with meta-analysis aimed to investigate the effects of recreational sports and combined training on BP and HbA1c in middle-aged and older adults.

## 2. METHODS

This review followed the Preferred Reporting Items for Systematic Reviews and Meta-analyses (PRISMA) guidelines [13] and was registered on the International Prospective Register of Systematic Reviews: PROSPERO (CRD42020207250)

### 2.1 Search Strategy

The electronic search was independently performed by two investigators (V.M.S and P.F) between July and August 2020 using the MEDLINE database (through PubMed), COCHRANE, and SciELO. For PubMed, Medical Subject Headings (MeSH) and Health Sciences Descriptors (DeCS) descriptors were combined using Boolean operators and the strategy is described in Electronic Supplementary Material (ESM) Table S1. Reference lists of original and review articles were also revised. There were no language restrictions, as well as on the publication date and there was no need to contact study authors to identify additional studies.

### 2.2 Eligibility Criteria

Articles describing randomized and nonrandomized clinical trials, conducted in men and women, aged ≥45 years with baseline systolic BP (SBP) ≥130mmHg or diastolic BP (DBP) ≥80 mmHg or type II diabetes would be eligible if they had evaluated recreational sports interventions or combined training for at least eight and no more than 52 weeks, and had reported baseline and endpoint measures of SBP, DBP and HbA1c.

To include combined training interventions, we selected those performing any aerobic exercise on land (i.e., cycling, walking, or running) simultaneously to resistance/strength training (aerobic and resistance exercises could be performed on the same day or on alternate days). Studies on recreational sports were included if the participants performed any team sport practice of a non-professional feature, with no complementary interventions. Studies conducted in participants who have restrains due to cardiac or pulmonary diseases, cognitive impairment, or studies that had co-interventions (diet, titration of medication) were excluded. We also excluded studies in which control groups performed exercise training.

### 2.3 Study Selection and Data Extraction

Two investigators (V.M.S and P.F) independently performed the screening of title and abstracts to identify potentially relevant articles. The searches were compared to assess agreement among investigator’s selection and the final sample was revised to exclude duplicated studies. Disagreements among searches were maintained to be solved in the next step. Subsequently, selected articles underwent a full-length reading and were independently analyzed following a standard criterion that determined both the inclusion and exclusion of studies. Disagreements regarding eligibility of the studies. were solved by consensus and, eventually, by a third party (R.F). The study selection process followed the PRISMA flowchart [14].

Data extraction was independently performed by two investigators (V.M.S and P.F), using a standardized form to obtain number of participants that were included in the analysis, sex, age, level of physical activity, BP and HbA1c values, adherence to the protocol (attend the sessions [%]) and participants dropouts [%]) and BP and HbA1c assessment techniques used in each study. Overall, the methods used to measure BP consisted of an automatic monitor or aneroid sphygmomanometer. For HbA1c, liquid chromatography, automated analyzer and immunoturbidimetric method were used in the measurement. Data were extracted in their original units to calculate summarized mean, standard deviation (SD) and mean difference before and after the training period for the outcomes SBP, DBP, HbA1c for both, recreational sports and combined training or control group. In instances where results were presented as mean ± SEM (mean standard error), SEM was converted to SD using SD = SEM × square root of populations (Sqrt^n).

### 2.4 Evaluation of Risk of Bias

Study quality assessment was conducted independently by two investigators (V.M.S and P.F), and disagreements were solved by consensus. The appraisal of methodological quality of the studies included the items proposed by Cochrane [15]: randomization methods, allocation concealment, blinding of patients and therapist, blinding of outcomes evaluators, incomplete outcomes, selective reporting of outcomes and other possible bias sources. The items were defined as high risk of bias, low risk of bias or unclear risk of bias.

### 2.5 Data Synthesis

Results are presented as the weighted mean difference (MD) for absolute values with a 95% confidence interval (CI) and using an inverse variance method with random effect models. Values of P <0.05 were considered statistically significant. Heterogeneity was assessed through Cochran Q (chi^2^) test, with *P* <0.05 indicating statistically significant heterogeneity between types of exercise effects among the individual trials. The extend of heterogeneity was quantified using the inconsistency index (*I*^*2*^) [16] and values >50% were considered to represent substantial heterogeneity. The analysis with all studies (exercise training [overall]) was performed considering the training modalities recreational sports and combined training used in the studies. Secondary analysis were conducted to test the influence on heterogeneity and estimated the effects of interventions according to SBP ≥130mmHg or DBP ≥80 mmHg and HbA1c ≥ 5,7% [18, 19] and characteristics of the intervention (training intensity and duration [weeks]). Statistical analyses were conducted using Review Manager, version 5.4 (Cochrane Collaboration).

## 3. RESULTS

### 3.1 Description of Studies

The electronic search identified 6002 titles and 15 additional articles were identified through reference lists. After excluding duplicates, articles were screened, with 245 underwent a full text review, and 27 studies were included in the systematic review and meta-analysis (Figure 1). The analysis included 1411 participants with age of 55.8 ± 8.3 years (range, 45–73 years) and 53% women. Across the sample, 9 out of 27 trials investigated recreational sports *vs*. no exercise (control group) with 125 and 104 participants, respectively, enrolled in soccer (n=7), rugby (n=1), and skiing (n=1). The remaining 18 studies evaluated combined training *vs*. no exercise, as a control group, including 654 and 526 participants, respectively. Table 1 shows the main characteristics of the studies.

**Table 1.** Sample characteristics of included studies

| Study | N | Age (years)<br>[mean ± SD] | Dropouts<br>(n) | Other characteristics |
| --- | --- | --- | --- | --- |
| Agner et al. 2018 [44] | 23 | 66 ± 4 | 0 | Sedentary, T2DM,<br>Hypertension |
| Amaro-Gahete et al. 2019 [45] | 17 | 54 ± 4. | 4 | Sedentary, Healthy |
| Andersen et al. 2010 [46] | 13 | 46 ± 7 | 1 | Sedentary, Hypertension |
| Andersen et al. 2014 [30] | 12 | 50 ± 7 | 0 | Sedentary, T2DM |
| Andersen et al. 2016 [47] | 9 | 68 ± 4 | 0 | Sedentary, Healthy |
| Balducci et al. 2004 [48] | 62 | 60 ± 8 | 10 | Sedentary, T2DM |
| Church et al. 2010 [49] | 76 | 55 ± 8 | 5 | Sedentary, T2DM |
| Da silva et al. 2020 [50] | 13 | 71 ± 4 | 0 | Sedentary, Healthy |
| Da silva et al. 2020 [50] | 13 | 63 ± 7 | 0 | Sedentary, Healthy |
| Dobrosielski et al. 2012 [51] | 51 | 57 ± 6 | 19 | Sedentary, T2DM |
| Jorge et al. 2011 [52] | 12 | 57 ± 9 | None | Sedentary, T2DM |
| Krustrup et al. 2017 [53] | 19 | 45 ± 6 | 3 | Sedentary, Hypertension |
| Lambers et al. 2008 [54] | 17 | 58 ± 9 | 2 | T2DM |
| Loimaala et al. 2003 [55] | 24 | 53 ± 6 | 0 | Sedentary, T2DM |
| Mendham et al. 2015 [33] | 10 | 46 ± 6 | 1 | Sedentary, T2DM |
| Miura et al. 2008 [56] | 29 | 69 ± 6 | 0 | Sedentary, Healthy |
| Miura et al. 2008 [56] | 25 | 69 ± 7 | 0 | Sedentary, Healthy |
| Miura et al. 2015 [57] | 45 | 73 ± 6 | 8 | Sedentary, Hypertension |
| Miura et al. 2015 [57] | 55 | 72 ± 7 | 3 | Sedentary, Healthy |
| Mohr et al. 2014 [58] | 21 | 45 ± 3 | 0 | Sedentary, Hypertension |

**Table 1.** (continued)
| Study | Participants | Age (years)<br>[mean ± SD] | Dropouts<br>(n) | Other population<br>characteristics |
| --- | --- | --- | --- | --- |
| Niederseer et al. 2011 [59] | 22 | 66 ± 2 | 5 | Sedentary, Healthy |
| Ohkubo et al. 2001 [60] | 32 | 67 | 0 | Sedentary, Healthy |
| Park et al. 2015 [61] | 24 | 71 ± 3 | 15 | Sedentary, T2DM,<br>Hypertension |
| Ruangthai et al. 2019 [62] | 16 | 67 ± 5 | 1 | Sedentary, Hypertension |
| Schmidt et al. 2013 [63] | 10 | 51 ± 7 | 4 | Sedentary, T2DM,<br>Hypertension |
| Schmidt et al. 2014 [64] | 9 | 68 ± 3 | 1 | Sedentary, Healthy |
| Shiotsu et al. 2018 [65] | 15 | 70 ± 3 | 0 | Sedentary, Hypertension |
| Shiotsu et al. 2018 [65] | 15 | 70 ± 3 | 0 | Sedentary, Hypertension |
| Stewart et al. 2005 [66] | 51 | 63 | 6 | Sedentary, Hypertension |
| Tessier et al. 2000 [67] | 19 | 69 ± 4 | 5 | Sedentary, T2DM |
| Timmons et al. 2018 [68] | 21 | 69 ± 2 | 0 | Sedentary, Healthy |

Values reported as mean ± standard deviation (SD); *T2DM* type II diabetes mellitus

**Fig. 1.**
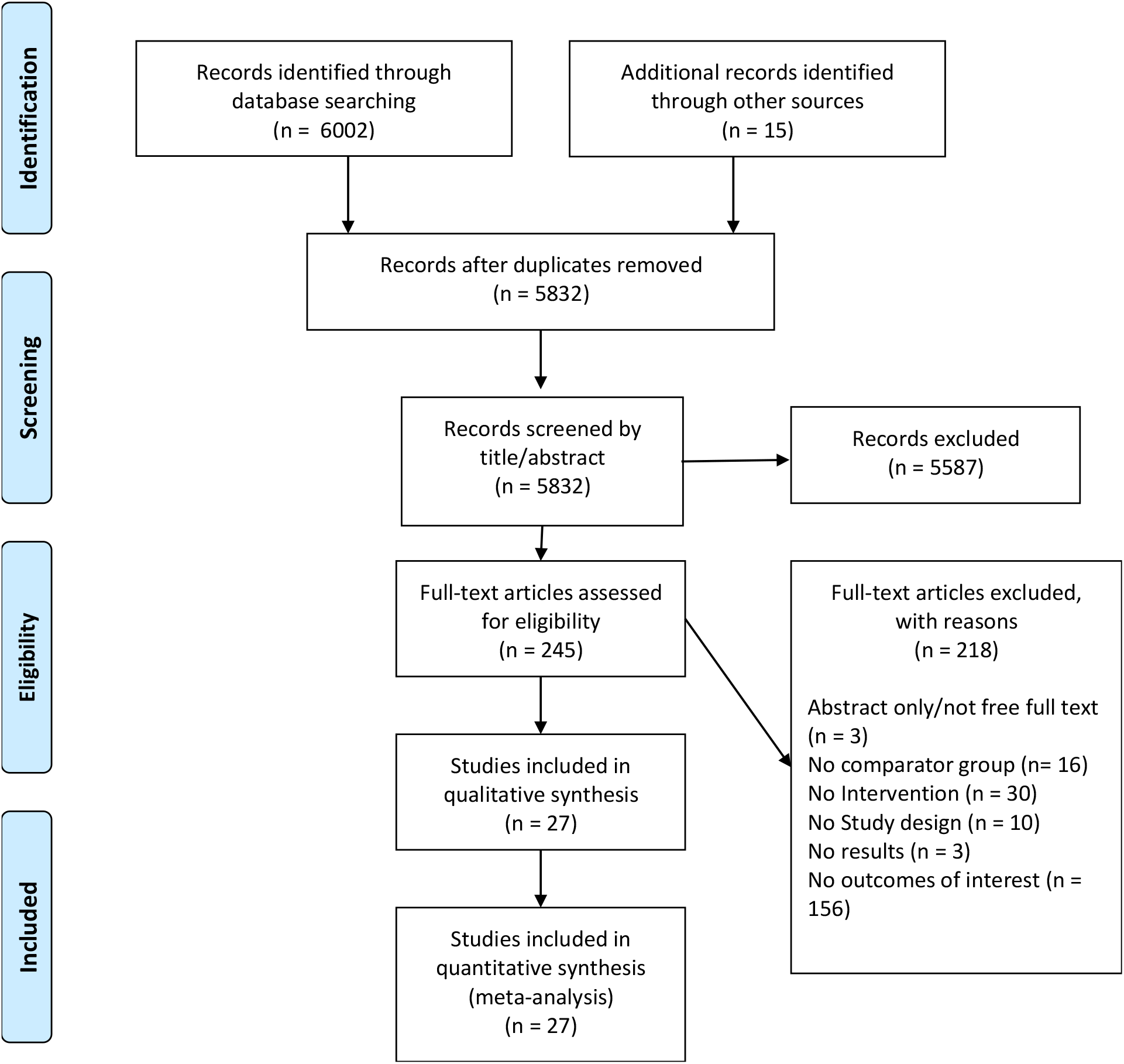
Flow diagram of the study selection process

Table 2 presents the characteristics of the exercise groups with all recreational sports interventions (9 studies) conducted for ≥ 8 weeks (range 8-52 weeks), five groups trained for ≥ 24 weeks and the participants trained for ≥ 2 days/week (range 2-3 days). The intensity of recreational sports interventions were classified as moderate (1 studies; <80% maximum heart rate [HRmax.]) or vigorous (6 studies; ≥80% HRmax.) according to ACSM guidelines [17], and two studies did not report exercise intensities. The sports sessions consisted of 60 minutes per day and 120 to 180 minutes per week. The attendance rate of recreational sports was 76 ± 13%.

**Table 2.**
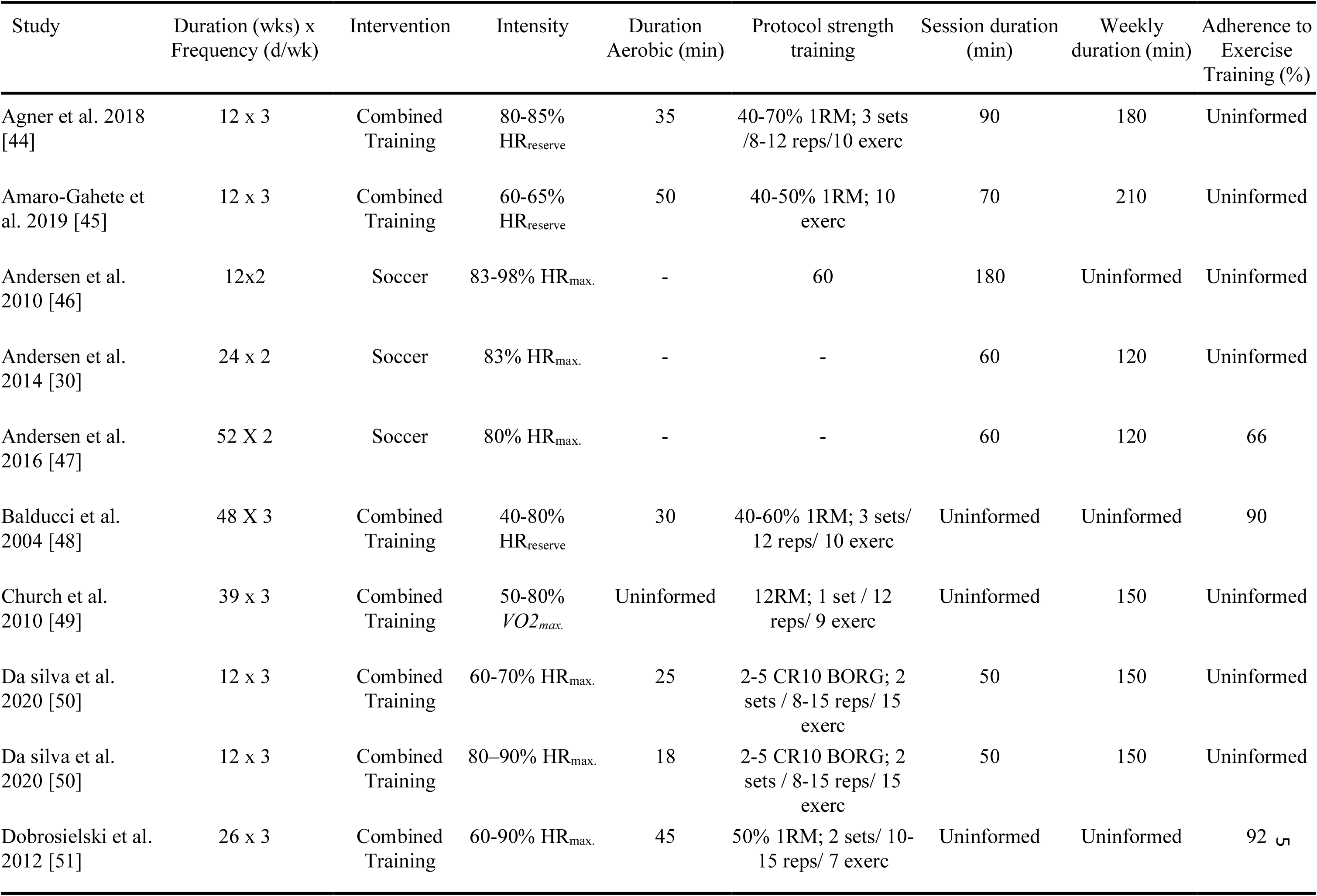
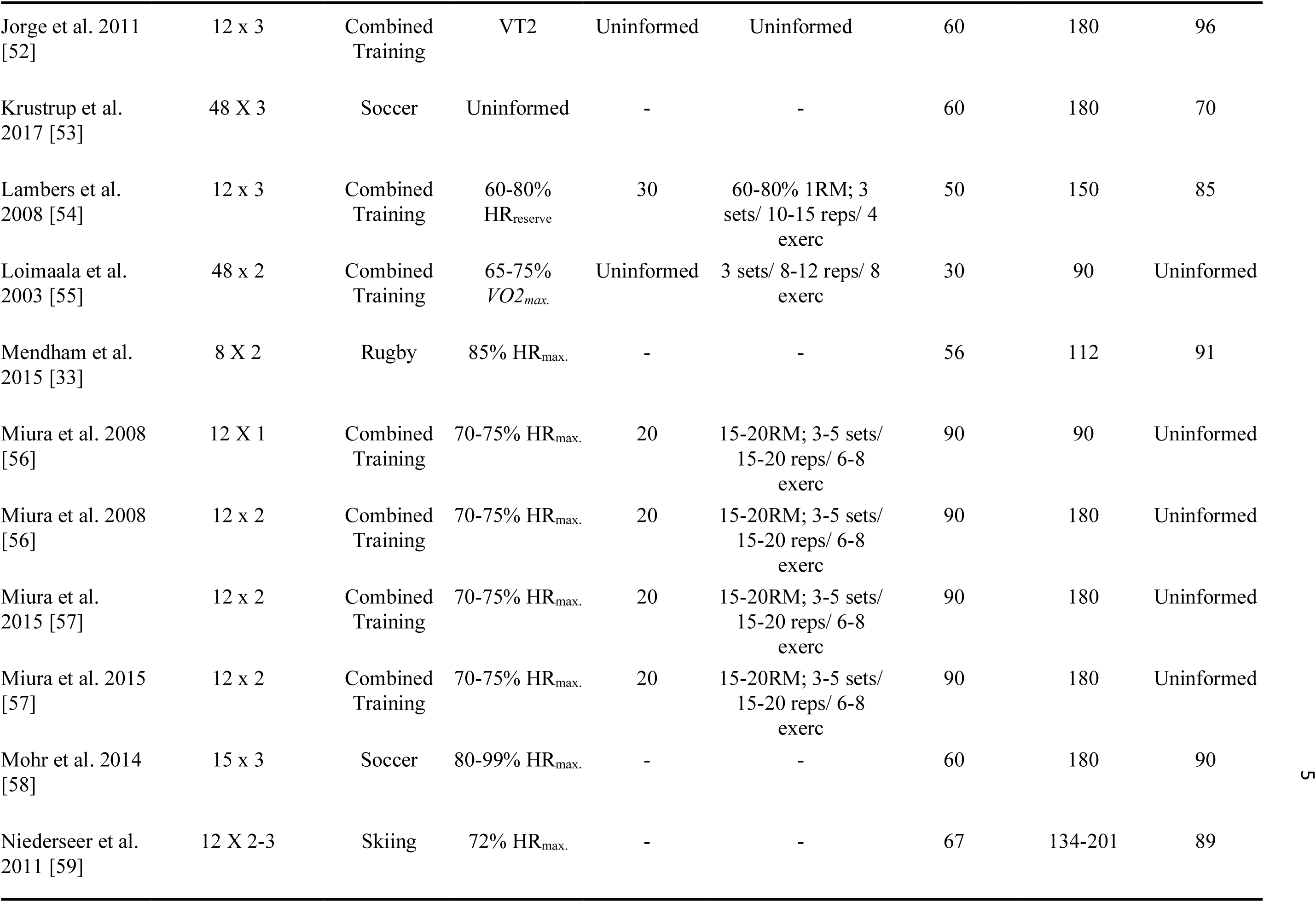
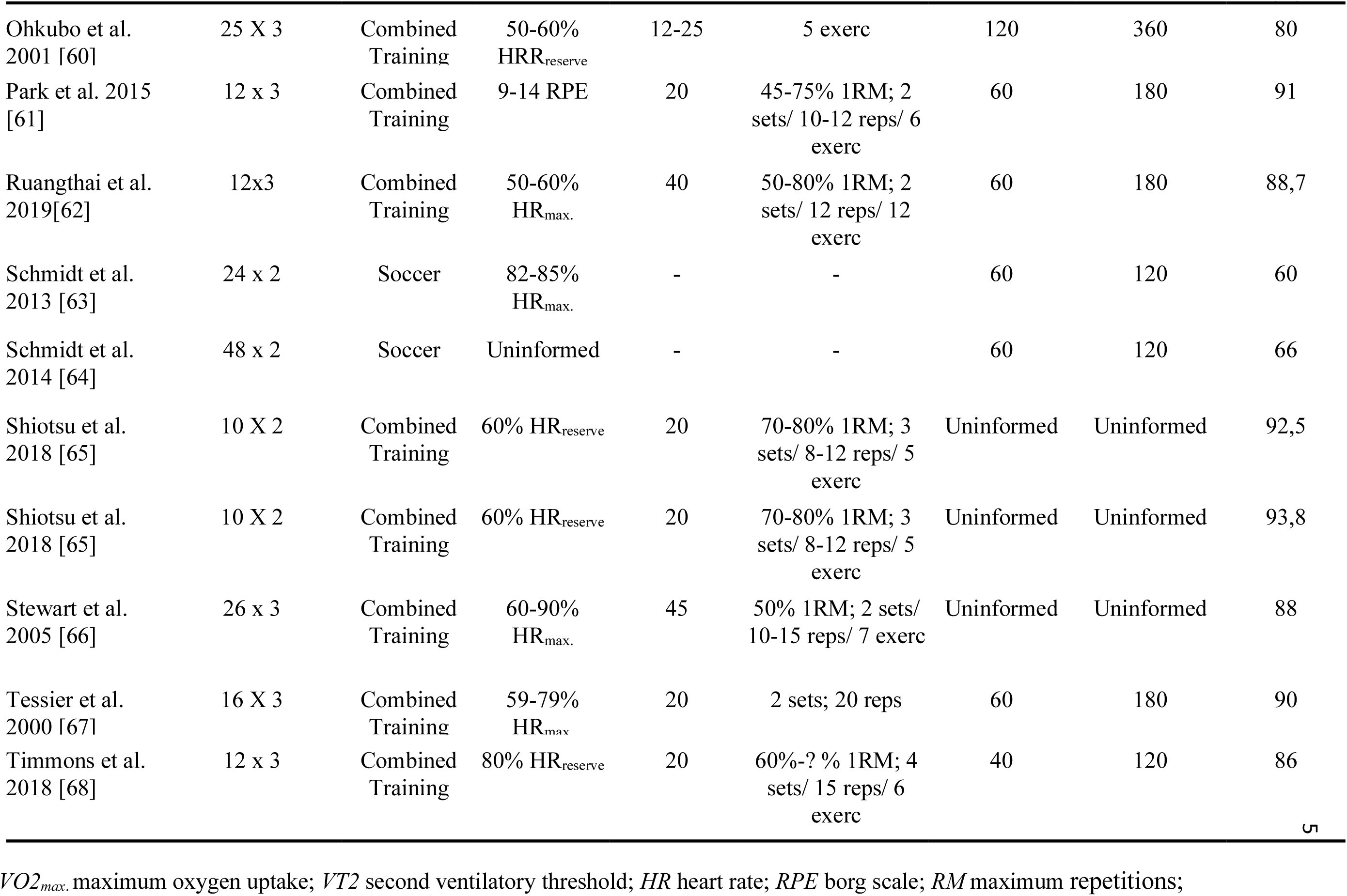
Characteristics of recreational sports and combined training interventions of included studies

Combined training interventions (18 studies) were conducted for ≥ 10 weeks (range 10-48 weeks) and participants trained for ≥1 days/week (range 1-4 days). Six groups trained for ≥ 24 weeks. Intensity of aerobic training was classified as moderate (16 studies; <80% HRmax.) or vigorous (2 studies; ≥80% HRmax.) [17]. The aerobic training was performed for 26.6 ± 9.8 minutes per session.

The intensity of strength training was carried out using percentages 60% (range 40-90%) of one-maximum repetition test (10 studies) or performed 15 (range 8-30 repetitions) maximum repetitions (3 studies) or in 5 (range 2-5) on the perception Borg scale CR-10 [18] (1 study), and four studies did not provide information. Overall, the strength training protocols included 2-3 sets per exercise, 6-30 repetitions, and a total of 4-10 exercises. The total duration of the sessions combined training consisted of 70 ± 26 minutes and a weekly range of 90 to 360 min. The attendance rate of combined training was 90 ± 4%.

### 3.2 Risk of bias assessment

Two reviewers evaluated methodological quality of the 27 studies included in this review, and 25 presented adequate generation of randomized sequence, 11 reported allocation concealment, 7 blinding of patients and therapist, 7 blinding of outcomes evaluators; 13 presented incomplete outcomes and 8 selective reports of outcomes. Eight studies presented unclear risk in other bias sources. Data on the risk of bias from studies individually are showed in ESM Figure S1.

### 3.3 Blood Pressure

An overall BP reduction, illustrated in Figures 2 and 3, to any exercise training vs. control group was found in SBP (MD: -4.3 mmHg; 95%CI: -6.1 to -2.5; *I*^*2*^ = 71%; *P* for heterogeneity <0.001) and DBP (MD: -3.0 mmHg; 95%CI: -3.7 to -2.3; *I*^*2*^ = 8%; *P* for heterogeneity = 0.36).

**Fig. 2.**
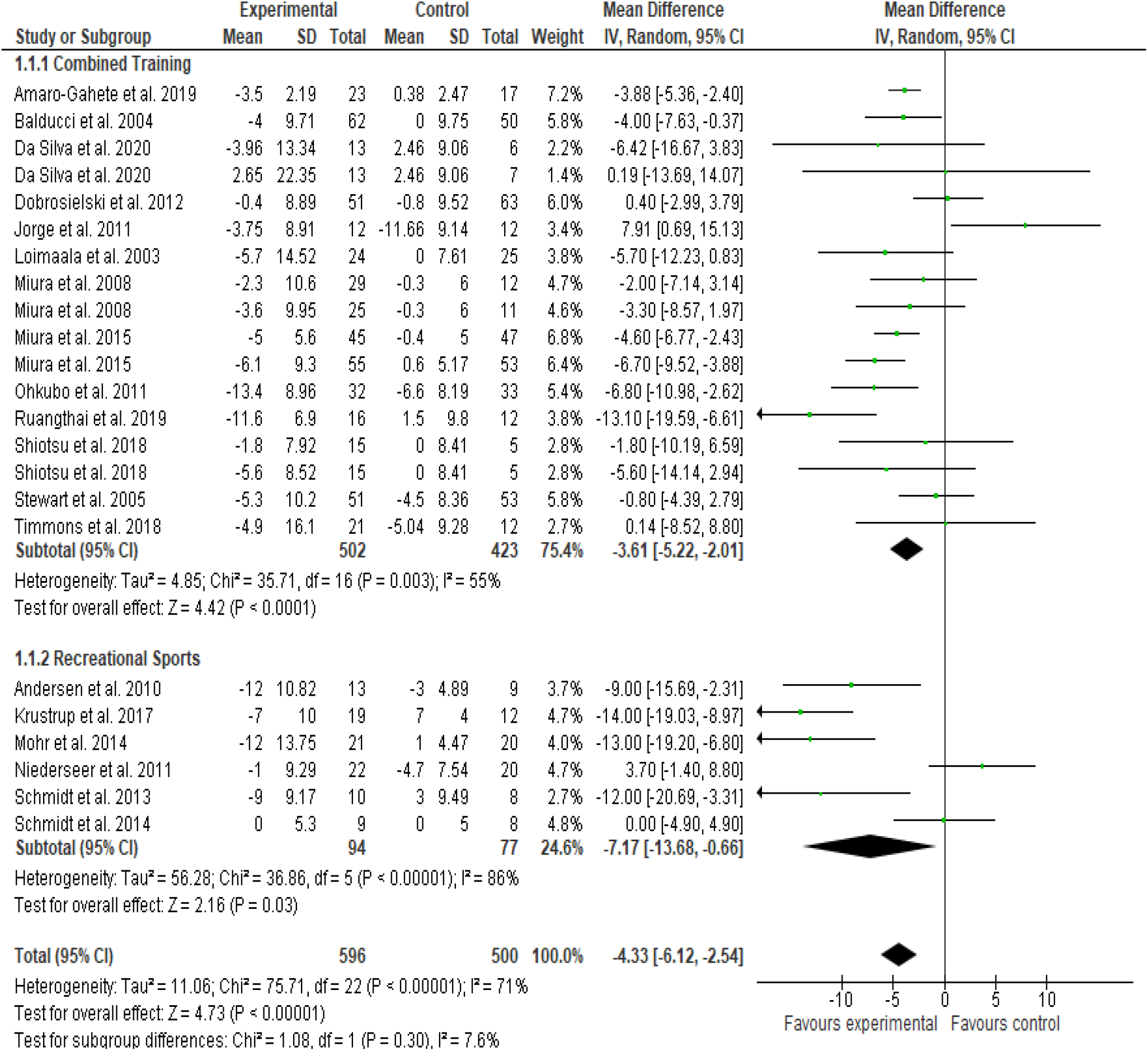
Absolute changes in systolic blood pressure (SBP) in individual studies of exercise training (overall) and subgroup analyses were carried out considering the training modalities: recreational sports and combined training. Squares represent study-specific estimates; diamonds represent pooled estimates obtained using a random-effect analyses

**Fig. 3.**
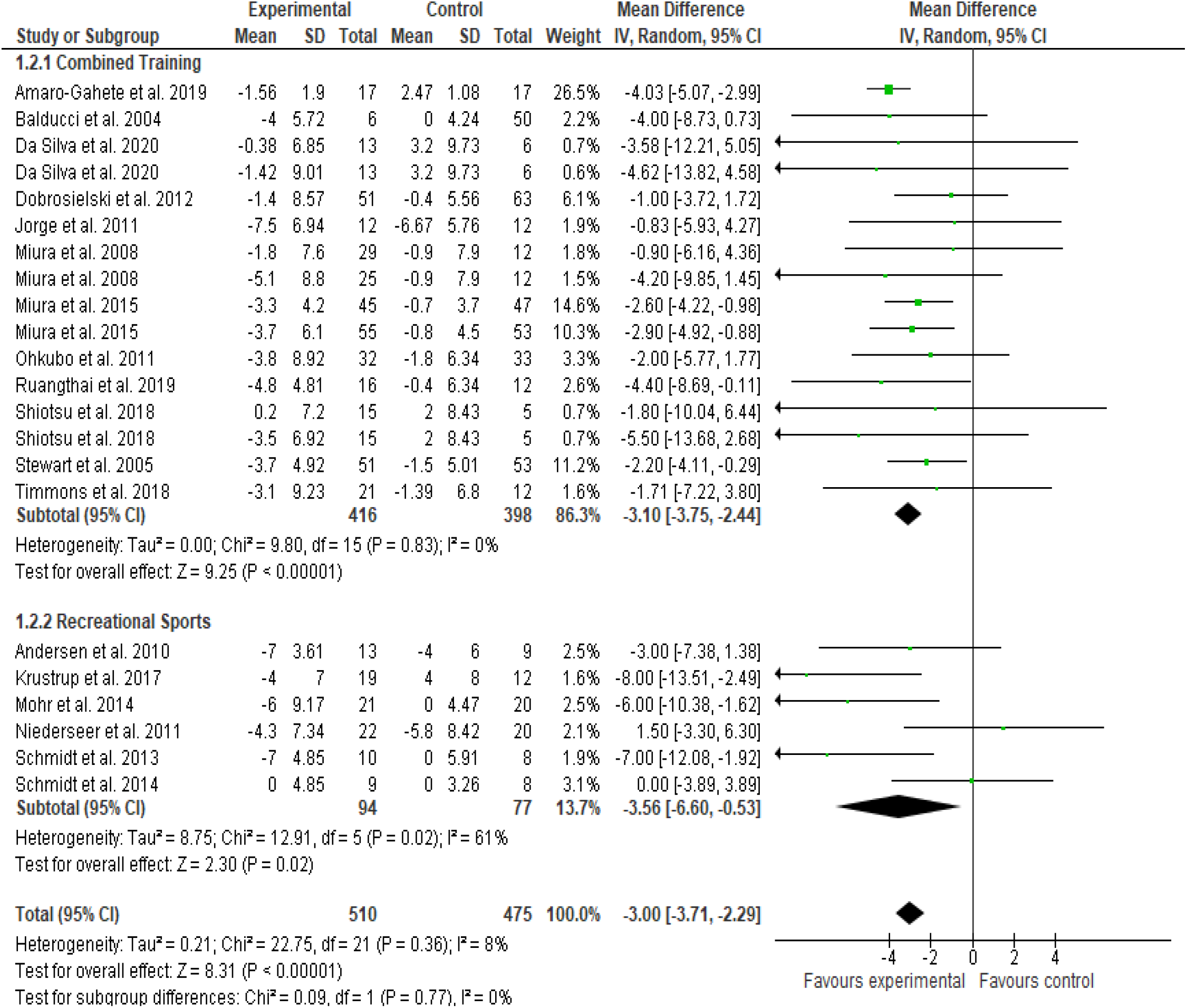
Absolute changes in diastolic blood pressure (DBP) in individual studies of exercise training (overall) and subgroup analyses were carried out considering the training modalities: recreational sports and combined training

The magnitude of SBP and DBP reduction after recreational sports and combined training programs is shown in Figures 2 and 3, respectively. Recreational sports (n=6 intervention groups, with 171 participants) were associated with a reduction in SBP (MD: -7.1 mmHg; 95%CI: -13.7 to -0.7; *I*^*2*^ *=* 86%; *P* for heterogeneity < 0.001) and DBP (MD: -3.5 mmHg; 95%CI: -6.6 to -0.5; *I*^*2*^ *=* 61%; *P* for heterogeneity = 0.02) compared with controls. Combined training (n=17 intervention groups, with 925 participants) was associated with a reduction in SBP (MD: -3.6 mmHg; 95%CI: -5.2 to -2.0; *I*^*2*^ *=* 55%; *P* for heterogeneity = 0.003) and DBP (n=16 intervention groups, with 814 participants) (MD: -3.1 mmHg; 95%CI: -3.7 to -2.4; *I*^*2*^ *=* 0%; *P* for heterogeneity = 0.83) compared with controls.

### 3.4 Glycosylated Hemoglobin (HbA1c)

The overall reduction in HbA1c is illustrated in Figure 4A for any exercise training vs. control group (MD: -0.40%; 95%CI: -0.56 to -0.23; *I*^*2*^ *=* 0%; *P* for heterogeneity = 0.57). The magnitude of HbA1c reduction after recreational sports and combined training programs is show in Figure 4B. Combined training (n=11 intervention groups, with 600 participants) was associated with reduction in HbA1c (MD: -0.47%; 95%CI: -0.67 to -0.27; *I*^*2*^ *=* 1%; *P* for heterogeneity = 0.43) compared with controls. No significant reduction (P= 0.12) was found in HbA1c for recreational sports (n=4 intervention groups, with 77 participants) (MD: -0.24; 95%CI: -0.53 to 0.06; *I*^*2*^ = 0%; *P* for heterogeneity = 0.87) compared with controls.

**Fig. 4.**
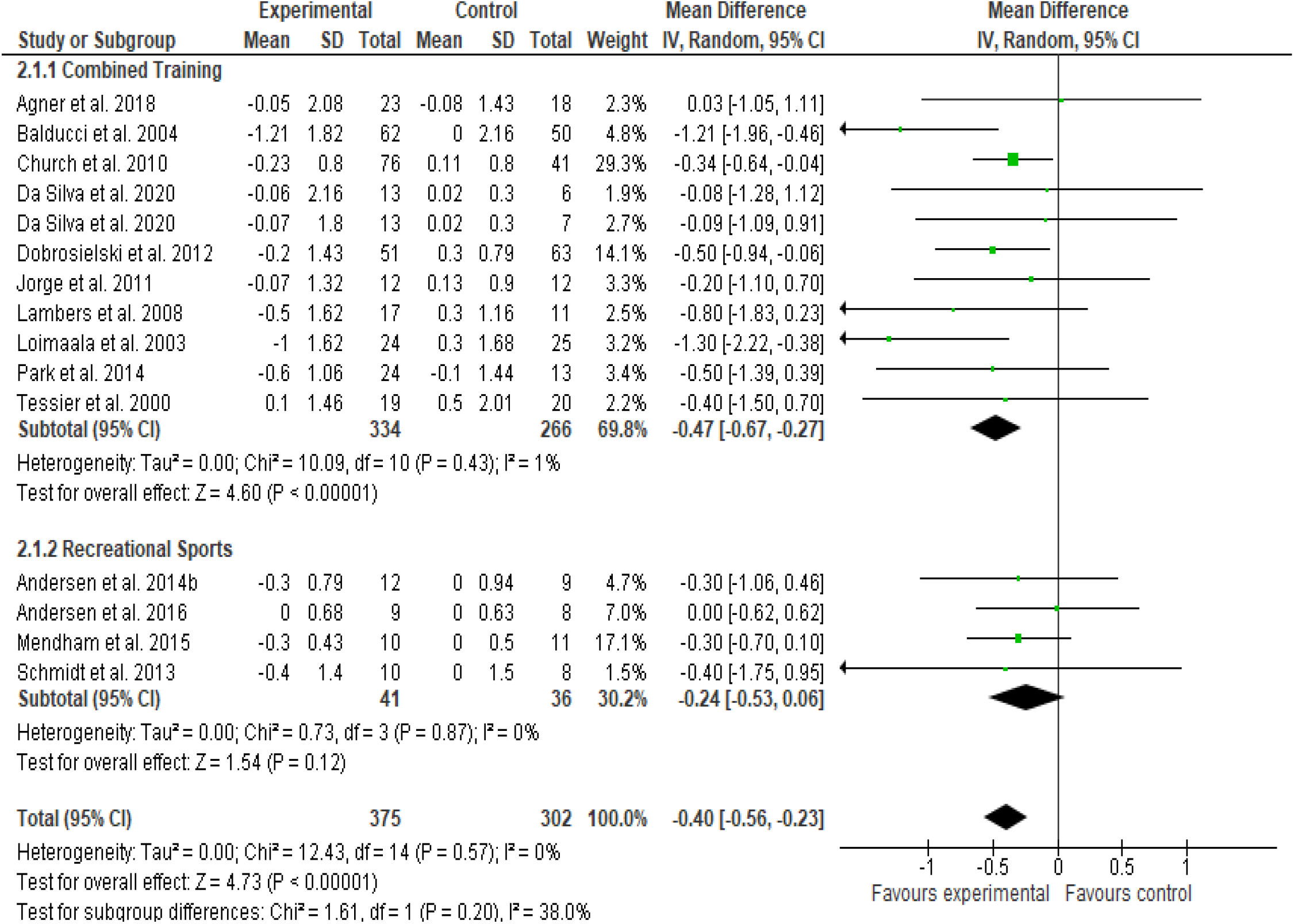
Absolute changes in glycosylated hemoglobin (HbA1c) in individual studies of exercise training (overall) and subgroup analyses were carried out considering the training modalities: recreational sports and combined training

### 3.5 Secondary Analyses and Exploration of Heterogeneity

Due to the high heterogeneity in studies involving interventions with combined training (*P* = 0.003) and recreational sports (*P* <0.001) in the SBP, DBP (only recreational sports), and the necessity to investigate the greatest benefits of these interventions in the above mentioned variables, secondary analyses were performed according to the baseline SBP and DBP values ≥130 or ≥80 mmHg, respectively, HbA1c ≥ 5.7% and we also carry out analyses according to the intensity and duration of the intervention.

#### Blood pressure

High baseline SBP and DBP values were associated with greater reduction (ESM, Table S3). Recreational sports-based interventions (n=4 intervention groups, with 112 participants) were associated with greater reduction in SBP (MD: -12.4 mmHg; 95%CI: -15.5 to -9.2; *I*^*2*^ *=* 0%; *P* for heterogeneity = 0.70) and DBP (MD: -5.7 mmHg; 95%CI: -8.1 to -3.3; *I*^*2*^ *=* 0%; *P* for heterogeneity = 0.49) compared with controls. Combined training (n=9 intervention groups, with 514 participants in SBP and n=8 intervention groups, with 344 participants in DBP) was associated with a reduction in SBP (MD: -3.9 mmHg; 95%CI: -6.7 to -1.2; *I*^*2*^ *=* 66%; *P* for heterogeneity = 0.003) and DBP (MD: -2.6 mmHg; 95%CI: -3.7 to -1.5; *I*^*2*^ *=* 0%; *P* for heterogeneity = 0.90) compared with controls.

Analyses according to the intensity of exercise and the duration of the interventions are presented in ESM Table S4. The results showed that recreational sports performed with higher intensities (n=5 intervention groups, with 129 participants) was associated with greater SBP reductions (MD: -9.4 mmHg; 95%CI: -15.3 to -3.5; *I*^*2*^ = 79%; *P* for heterogeneity = 0.009), and DBP (MD: -4.5 mmHg; 95%CI: -7.5 to -1.5; *I*^*2*^ = 52%; *P* for heterogeneity = 0.08) compared with controls. Analyzing the interventions duration, recreational sports lasting ≥ 24 weeks (n=3 intervention groups, with 59 participants) showed no significance reduction in SBP (MD: -8.5 mmHg; 95%CI: -18.3 to 1.4; *I*^*2*^ = 88%; *P* for heterogeneity = 0.003), and studies for < 24 weeks (n=2 intervention groups, with 83 participants) did not significantly reduce SBP (MD: -4.5 mmHg; 95%CI: -20.9 to 11.8; *I*^*2*^ = 94%; *P* for heterogeneity < 0.001) compared with controls. On the other hand, for DBP, recreational sports lasting ≥ 24 weeks (n=3 intervention groups, with 88 participants) was associated reduction in DBP (MD: -4.2 mmHg; 95%CI; -7.9 to -0.5; *I*^*2*^ = 60%; *P* for heterogeneity = 0.06) and studies lasting < 24 weeks (n=2 intervention groups, with 83 participants) did not reduced DBP (MD: -2.3 mmHg; 95%CI: -9.7 to 5.0; *I*^*2*^ = 80%; *P* for heterogeneity = 0.02) compared with controls.

Secondary analyses were conducted in interventions using combined training. No significant SBP reduction was found with combined training performed with higher intensities (n=3 intervention groups, with 238 participants) (MD: -0.1 mmHg; 95%CI: -2.6 to 2.3; *I*^*2*^ = 0%; *P* for heterogeneity = 0.89) and lower intensities reduction SBP (n=14 intervention groups, with 687 participants) (MD: -4.4; 95%CI: -6.0 to -2.7; *I*^*2*^ = 48%; *P* for heterogeneity = 0.02) compared with controls. For DBP, similar reductions were detected for combined training, performed at higher intensities (n=3 intervention groups, with 238 participants) (MD: -1.9 mmHg; 95%CI: -3.4 to -0.3; *I*^*2*^ = 0%; *P* for heterogeneity = 0.65) and lower intensities (n=13 intervention groups, with 577 participants) (MD: -3.4; 95%CI: -4.1 to -2.6; *I*^*2*^ = 0%; *P* for heterogeneity = 0.91) compared with controls.

According to the duration of interventions, combined training duration ≥ 24 weeks (n=5 interventions, 444 participants) was associated with a reduction in SBP (MD: -3.0; 95%CI: - 5.8 to -0.2; *I*^*2*^ = 57%; *P* for heterogeneity = 0.05) and studies lasting < 24 weeks (n=12 interventions, 475 participants) improvement reduction in SBP (MD: -3.9; 95%CI: -5.9 to -1.9; *I*^*2*^ = 54%; P for heterogeneity = 0.01) compared with controls. In DBP, combined training consisting of studies with ≥ 24 weeks (n=4 interventions, 339 participants) was associated a reduction (MD: -2.0; 95%CI: -3.4 to -0.6; *I*^*2*^ = 0%; *P* for heterogeneity = 0.74) and also studies with < 24 weeks (twelve interventions, 475, participants) reduced DBP (MD: -3.4; 95%CI: -4.1 to -2.66; *I*^*2*^ = 0%; *P* for heterogeneity = 0.90) compared with controls.

#### Glycosylated Hemoglobin

The combined training, all participants had an average baseline HbA1c values ≥5.7% and no heterogeneity was identified. Therefore, no secondary analyses were performed with this variable. During the sport interventions, in 3 of 4 interventions (60 participants) the participants presented mean HbA1c values ≥5.7%. No significant reduction in absolute HbA1c (MD: -0.31%; CI: -0.65 to 0.03; *I*^*2*^ *=* 0%; *P* for heterogeneity = 0.99) was found after recreational sports compared with controls (ESM Table S3).

Analyses according to the diagnosis of hypertension at baseline for SBP values ≥130mmHg, were only able to identify the high heterogeneity of SBP in interventions with recreational sports.

## 4. DISCUSSION

Blood pressure and glycosylated hemoglobin control are key components to prevent and treat hypertension and type 2 diabetes, two chronic conditions with increased prevalence and severity throughout lifespan [19-21]. Besides, they are also predictors of increased mortality [22, 1]. To the best of our knowledge, this is the first meta-analysis that assessed the effects of recreational sports and combined training on BP and HbA1c in middle-aged and older adults. We found that both exercise interventions reduced BP compared with control (Recreational sports: systolic/diastolic BP - 7/4 mmHg; Combined training: systolic/diastolic BP - 4/3 mmHg). Besides, combined training also reduced HbA1 compared to the control group (0.47%), and there was a trend toward reduction after recreational sports, found in participants with baseline HbA1c ≥5.7% (−0.31%; *P* = 0.08). In our analyses, recreational sports and combined training were not directly compared because no original study using combined training and recreational sports interventions is available. The present meta-analysis confirmed the efficacy of combined training to improve BP and HbA1c control and provided evidence to support that recreational sport should be considered an interesting alternative to traditional exercises to improve these cardiometabolic parameters, especially BP, in middle-aged and older adults.

Sustained elevation of BP represents a marked condition from the fourth decade of life onwards [21], and physical exercises should be recommended as a key lifestyle therapy for adults with high blood pressure (BP) for the prevention, treatment, and control of hypertension [20]. Effective strategies for BP control are associated to reduction of up to 50% of cardiovascular events [1] with a linear association between the magnitude of BP reduction and the risk of cardiovascular disease and all-cause mortality [23]. Our findings have suggested important reductions on BP after recreational sports (systolic/diastolic BP - 7/4 mmHg) and combined training (systolic/diastolic BP - 4/3 mmHg) interventions. Considering the above-mentioned associations, this reduction of 4-7 mmHg in systolic BP can reduce cardiac morbidity by 5%, stroke by 8-14%, and all-cause mortality by 4% [20, 24]. In addition, our secondary analyses suggested that hypertensive individuals are prone to achieve greater magnitudes of systolic and diastolic BP reduction only after recreational sports (systolic/diastolic BP - 12/4 mmHg). This result suggests that recreational sports have the potential to be used as an effective strategy to reduce blood pressure in aging adults with hypertension.

Currently, data regarding the optimal exercise intensity to reduce BP remains controversial [25, 26]. We compared the effects of recreational sports and combined training performed at different intensities (i.e., low [<80% HRmax] and high [≥80% HRmax]). Recreational sports performed at higher intensities (systolic / diastolic BP - 9/4 mmHg) and combined training performed at lower intensities (systolic / diastolic BP - 4/3 mmHg) showed meaningful reductions in BP. The results suggest that there is a dose-response relationship, especially related to the intensity of exercise in recreational sports and combined training. We observed that recreational sports performed at higher intensities promoted greater magnitudes of reduction in BP than with combined training performed in lower intensities. Something that makes us believe that moderate to higher intensities should be considered to cause great magnitudes of reduction in BP [25, 27]. In contrast, the combined training performed at higher intensities did not show a significant reduction for SBP (−0.1 mmHg; P = 0.90), but for DBP (−2 mmHg; P = 0.02). However, the combined training performed at higher intensities was performed gradually, with the intensity progression occurring over weeks, starting at a lower intensities (<50% HR_max_) in weeks 1-3, until reaching high intensity (≥80% HR_max_) in the final weeks of training, something that possibly interfered with the results when stratifying the combined training at high intensity.

In this systematic review and meta-analysis, combined training has demonstrated statistically significant reduction in HbA1c compared to the control group (−0.47%) This finding is in accordance with a previous meta-analysis [4, 29], confirming the benefits of combined training on glycemic management. An important difference from the previous meta-analysis was that we did not include interventions that combine exercise with dietary monitoring or medication titration, highlighting the benefits of exercise alone to reduce HbA1c. In addition, participants were aged >45 years, providing concise evidence for those who are at risk of developing type 2 diabetes [19]. In relation of the benefits of recreational sports on HbA1c, we have found a trend toward reduction in HbA1c compared to the control group in participants with HbA1 ≥5.7%. Although this result is promising, future studies are necessary to confirm the effectiveness of recreational sports to reduce HbA1c. Few interventional studies assessed the benefits of recreational sports, resulting in lower number of participants included in our analysis. If compared to the number of studies included in the combined training analyses, we have limited statistical power that may have affected this result [30]. Besides, the baseline HbA1c values of participants of recreational sports that were included in our analyses were lower than those that performed combined training, and the magnitude of glycemic reduction after exercise is directly related to the baseline values of participants [31]. Future randomized clinical trials of high methodological quality should be carried out to confirm the benefits of recreational sports on HbA1c in aging adults.

Exercise adherence plays a vital role in maximizing the desired benefits of exercise, since continuity is essential to maintain the broad-spectrum health improvements resulted from exercise practice. Different studies have observed a high rate of session attendance during recreational sports compared to traditional exercise programs [32, 33]. The monotony and reduced motivation that conventional physical exercise programs may provide [34, 35], and the positive psychosocial results and motivational aspects resulted from recreational sports [10] are potential explanations that could result in a lower exercise adherence after combined training. The results of our meta-analysis did not confirm these observations. The recreational sports studies included in the present review have reported an overall adherence of 76 %, and most of the trials lasted < 24 week. Of the 20 combined training studies included, 12 reported adherences to the intervention with an overall adherence over 90%. Thus, both interventions seem to be good strategies to improve adherence to exercise programs. An important factor that promotes adherence is the effectiveness of the intervention, since it positively impacts on motivation, confidence, and maintenance of the training program [36]. In the present review, both recreational sports and combined training lead to a good adherence and seems to be effective in improving health outcomes [37]. We emphasize that future studies should directly compare the two forms of exercise to better understand the factors that lead to adherence to these interventions.

Adults with diabetes are at high risk of developing hypertension because common aspects of pathophysiology are shared by both conditions [38]. Studies have demonstrated associations of hyperglycemia with endothelial dysfunction, vascular stiffness [39], reduced nitric oxide production [40], and blood viscosity [41], which are considered key mechanisms linked to increased hypertension and cardiovascular disease [39]. Diabetes and hypertension often coincide, so strategies aimed at improving glucose control may reduce the risk of hypertension among people with diabetes [42]. On the other hand, a small reduction in SBP and DBP has been associated with a reduction in cardiovascular events and mortality in the general population [1] and in patients with diabetes [43]. And even reductions in BP within the normal range can be clinically significant, especially among aging adults [1]. In this context, the results of the present review reinforce the idea of using physical exercise as a non-pharmacological therapy that can help the management of diabetes and hypertension.

This systematic review has some limitations that must be considered. Overall, general quality of the studies was low, reflecting an increased risk of bias since a significant part of the studies included in our analyses did not provide an adequate detailed description of the methods. Other possible limitation is related to the method used to assess BP. Even though office BP can be used to assess and diagnose hypertension, 24-h ambulatory BP monitoring is the gold standard for assessing BP behavior, and future research on the use of recreational sports and combined training should be assessed BP throughout long periods and under usual conditions. Our meta-analysis has strengths. Eligibility criteria that included only exercise interventions in middle-aged and older adults, without any dietary intervention or titration of medication, specifically highlighting the benefits of combined training and recreational sports for this population. It brings important implications for the exercise prescription targeted to aging individuals who have essential hypertension and/or type 2 diabetes. Combined training programs should be emphasized in older population, since this type of exercise is extremely effective in reducing HbA1c and BP. Recreational sports also proved to be a very effective exercise strategy to reduce BP, but still need additional randomized clinical trials to confirm its effectiveness in reducing HbA1c in middle-aged and older adults.

## CONCLUSION

Recreational sports and combined training are effective exercise interventions to reduce blood pressure in middle-aged and older adults. Additionally, combined training seems to be an excellent strategy to reduce HbA1c, and future studies are necessary to confirm the effectiveness of recreational sports to improve glycemic profile in aging population.

## Data Availability

The datasets generated and/or analyzed during the current study are available from the corresponding author on reasonable request.

---

**Electronic Supplementary Material Table S1.**
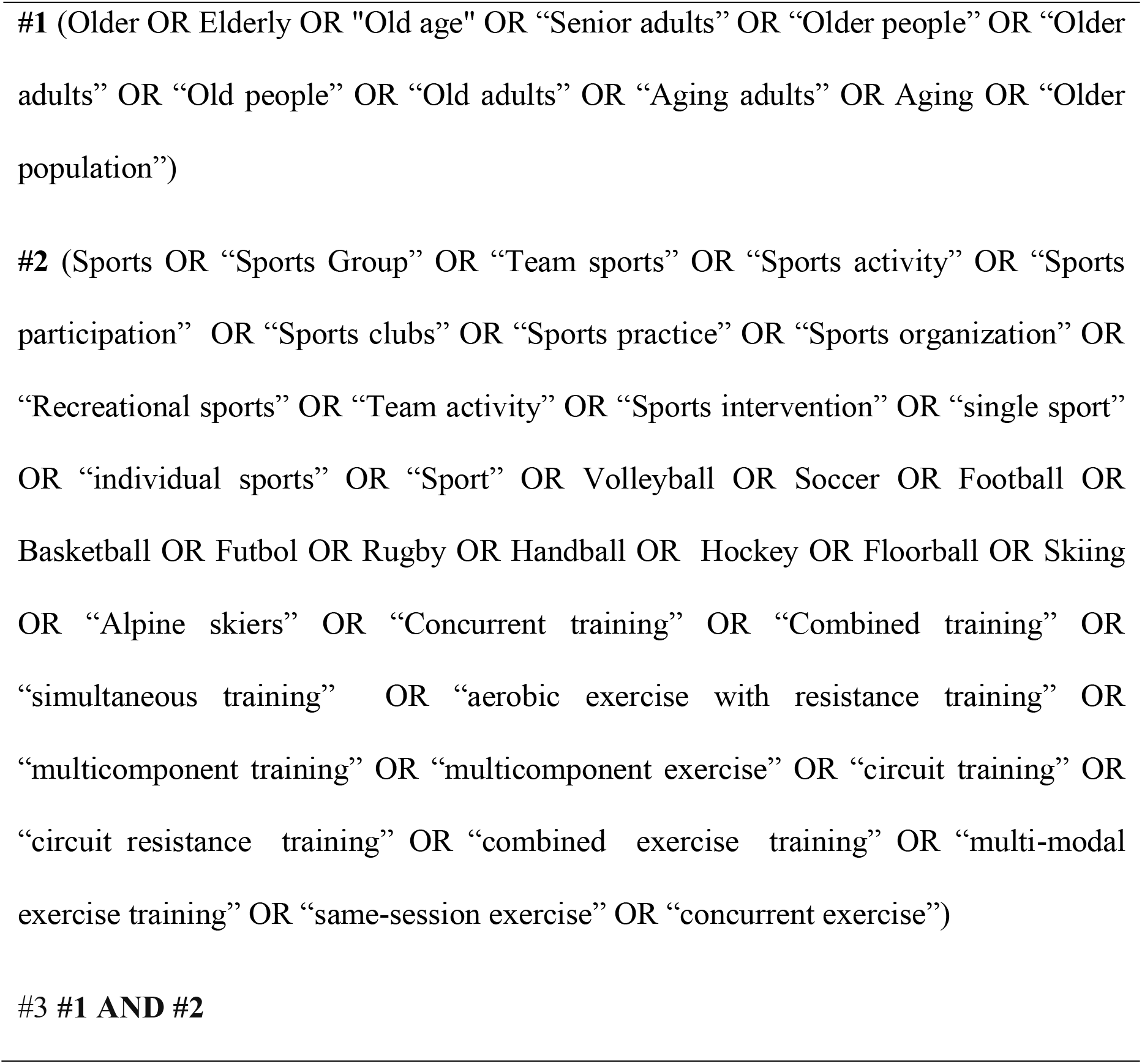
Literature search strategy used for the PubMed database

**Electronic Supplementary Material Figure S1.**
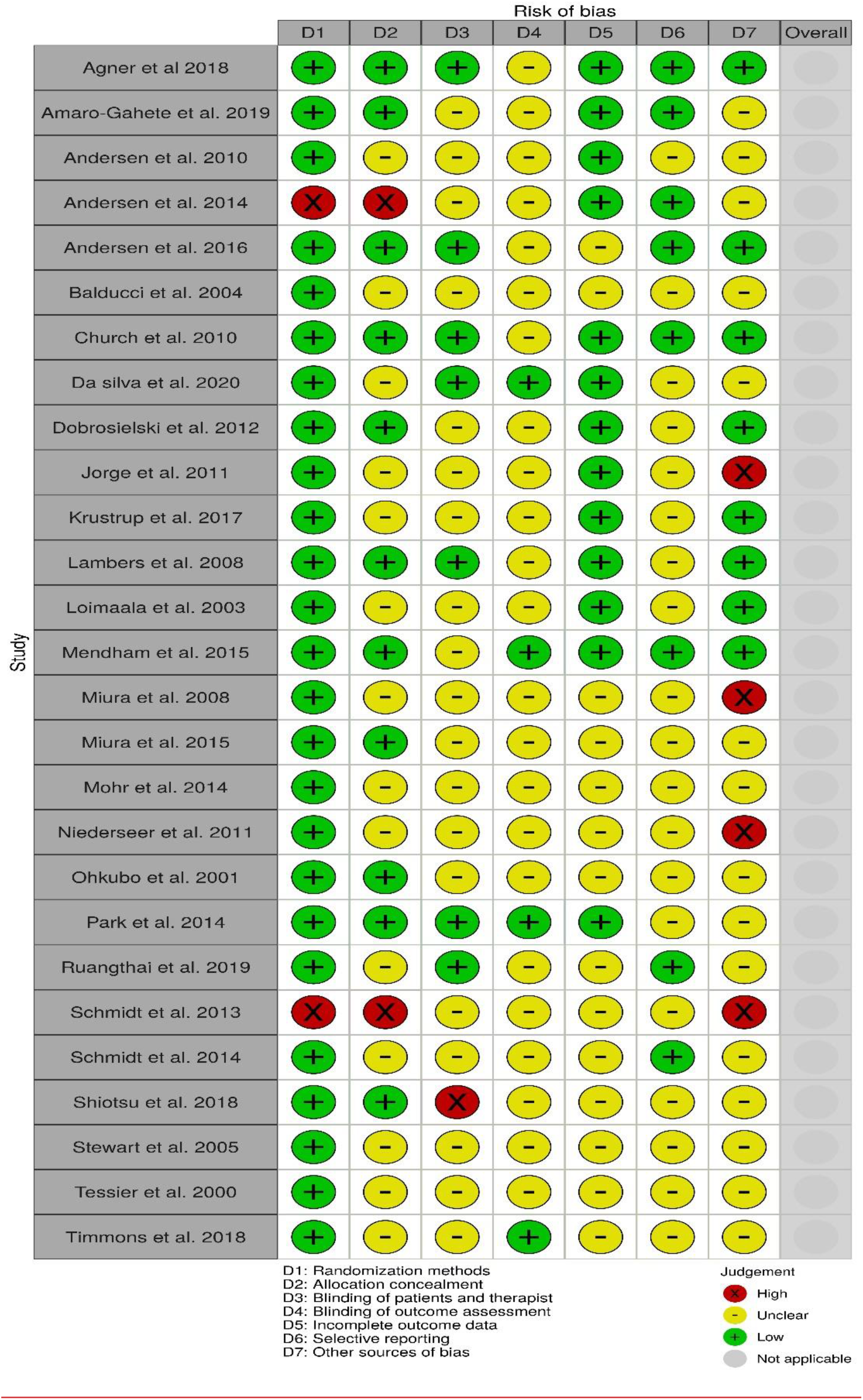
Risk of bias of studies included in the meta-analysis.

**Electronic Supplementary Material Table S3.** Absolute changes in systolic and diastolic blood pressure and glycosylated hemoglobin (HbA1c) in studies of recreational sports and combined training *vs*. no intervention stratified according to values according to SBP ≥130mmHg or DBP ≥80 mmHg and HbA1c ≥ 5,7%

| **Participants with BP $\geq 130/80$ mmHg** |  |  |  |
| --- | --- | --- | --- |
|  | **N** | **Mean Difference**<br>**(95%)** | **P** |
| **Recreational sports** |  |  |  |
| Systolic BP (mmHg) | 112 | -12.38 (-15.52; -9.23) | < 0.01 |
| Diastolic BP (mmHg) | 112 | -5.71 (-2.88; 0.18) | < 0.01 |
| **Combined training** |  |  |  |
| Systolic BP (mmHg) | 514 | -3.99 (-6.74; -1.23) | < 0.05 |
| Diastolic BP (mmHg) | 344 | -2.62 (-3.72; -1.51) | < 0.01 |
| **Participants with HbA1c $\geq 5,7\%$** |  |  |  |
|  | **N** | **Change (95% CI)** | **P** |
| **Recreational Sports** |  |  |  |
| HbA1c (%) | 60 | -0.31 (-0.65; 0.03) | 0.08 |
| **Combined Training** |  |  |  |
| HbA1c (%) | 600 | -0.41 (-0.67; -0.27) | < 0.01 |

P values statistically significant; *BP* blood pressure

**Electronic Supplementary Material Table S4.** Secondary analyses by intensity and intervention duration in absolute changes in systolic blood pressure and diastolic blood pressure in recreational sports and combined training

| **Subgroup** | **N of studies**<br>**(participant)** | **Mean Difference**<br>**(95%CI)** | **P** | ***I*<sup>2</sup>** | ***P*** |
| --- | --- | --- | --- | --- | --- |
| **Systolic BP – Intensity** |  |  |  |  |  |
| **Recreational sports** |  |  |  |  |  |
| High intensity (≥80% HRmax) | 5 (129) | -9.41 (-15.31; - 3.52) | .002 | 79% | < 0.09 |
| **Combined training** |  |  |  |  |  |
| Low intensity | 14 (687) | -4.40 (-6.04; - 2.75) | <.001 | 48% | 0.02 |
| High intensity | 3 (238) | -0.15 (-2.58; 2.27) | .90 | 0% | 0.89 |
| **Diastolic BP - Intensity** |  |  |  |  |  |
| **Recreational sports** |  |  |  |  |  |
| High intensity | 5 (129) | -4.49 (-7.47; -1.51) | .003 | 52% | 0.08 |
| **Combined training** |  |  |  |  |  |
| Low intensity | 13 (557) | -3.36 (-4.09; -2.64) | <.001 | 0% | 0.91 |
| High intensity | 3 (238) | -1.88 (-3.47; -0.34) | .02 | 0% | 0.65 |
| **Systolic BP– Intervention duration (weeks)** |  |  |  |  |  |
| **Recreational sports** |  |  |  |  |  |
| < 24 | 2 (83) | -4.55 (-20.82; 11.81) | .59 | 94% | < 0.01 |
| ≥ 24 | 3 (88) | -8.47 (-18.31; 1.37) | .09 | 88% | < 0.03 |
| **Combined training** |  |  |  |  |  |
| < 24 | 12 (475) | -3.96 (-5.98; -1.94) | <.001 | 54% | 0.01 |
| ≥ 24 | 5 (444) | -3.01 (-5.78; -0.24) | .03 | 57% | 0.05 |

| **Diastolic BP – Intervention duration (weeks)** |  |  |  |  |  |
| --- | --- | --- | --- | --- | --- |
| **Recreational sports** |  |  |  |  |  |
| < 24 | 2 (83) | -2.32 (-9.66; 5.03) | .54 | 80% | 0.02 |
| ≥ 24 | 3 (88) | - 4.16 (-7.86; 0.47) | .03 | 60% | 0.06 |
| **Combined training** |  |  |  |  |  |
| < 24 | 12 (475) | -3.41 (-4.15; -2.66) | <.001 | 0% | 0.90 |
| ≥ 24 | 4 (339) | -2.02 (-3.40; -0.64) | .004 | 0% | 0.74 |

Intensities were determined as Low (HRmax or 1-RM <80%) and High (HRmax & 1-RM ≥80%). *BP* blood pressure; *HR* heart rate; *RM* maximum repetitions. P values statistically significant; *P* values statistically significant for heterogeneity.

